# Relationship of subjective and objective cognition with post-stroke mood differs between early and long-term stroke

**DOI:** 10.1101/2024.01.08.24300985

**Authors:** Andrea Kusec, the OX-CHRONIC Team, Nele Demeyere

**Affiliations:** Nuffield Department of Clinical Neurosciences, University of Oxford, John Radcliffe Hospital, Oxford, OX3 9DU

**Keywords:** stroke, post-stroke depression, post-stroke anxiety, cognition, cognitive impairment, longitudinal cohort

## Abstract

**Background:** Depression and anxiety affects 1 in 3 stroke survivors. Performance on standardized objective cognitive tests and self-reported subjective cognitive complaints are associated with concurrent depression and anxiety, but it is unknown whether and how objective and subjective cognition relate to longer-term emotional outcomes.

**Method:** *N =* 99 stroke survivors (*M* age = 68.9, *SD* = 13.1; Median NIHSS = 5) from the OX-CHRONIC cohort completed measures of depression and anxiety (Hospital Anxiety and Depression Scale; HADS), objective cognition (Oxford Cognitive Screen) and subjective cognitive complaints (Cognitive Failures Questionnaire) at 6-months (Time 1), at ∼4.5 years (Time 2) and ∼5.5 years (Time 3) post-stroke. The contribution of objective and subjective cognition to depression and anxiety was determined via mixed-effects models.

**Results:** We found no evidence that age, stroke severity, years of education, and participant sex related to changes in HADS-Depression or HADS-Anxiety scores. Objective cognitive impairments at Time 1 (*b =* -0.79, *p* < .05) and increases in subjective cognitive complaints at Time 3 (*b =* 0.77, *p <* .05) related to increased HADS-Depression scores (Marginal R^2^ = 0.22). Only increases in subjective cognitive complaints at Time 3 (*b =* 0.96, *p <* .05) related to increased HADS-Anxiety scores (Marginal R^2^ = 0.20). When conducting models in reverse, HADS-Depression and HADS-Anxiety scores did not reciprocally explain changes in subjective cognitive complaints.

**Conclusions:** Objective cognitive abilities are more strongly associated with depression at 6-months post-stroke, while subjective cognitive complaints are more relevant to both long-term post-stroke depression and anxiety.

## Introduction

Stroke is a leading cause of mortality worldwide and increases risk of chronic disability in physical, psychological, and functional domains (Feigin et al., 2014). The impact of stroke often causes emotional distress, with post-stroke depression and anxiety prevalence rates nearly double the general population (Schottke & Giabbiconi, 2015; Cumming et al., 2016). Elevated depression and anxiety can persist for at least up to 10 years post-stroke (Ayerbe et al., 2014). Given this, investigating causes of post-stroke depression and anxiety has been ranked as the number one long-term research priority by stroke survivors, carers, and clinicians (Hill et al., 2022).

In addition to mood disorders, stroke commonly causes cognitive impairment, such as attention, memory, language, and executive function difficulties. Stroke survivors often experience multi-domain cognitive impairments (Lesniak et al., 2008; Demeyere et al., 2016; 2019) that can negatively affect everyday activities and participation (Nys et al., 2005; Bickerton et al., 2015; Demeyere et al., 2019; Mole & Demeyere 2020). Performance on objective, neuropsychological cognitive tasks has been associated with post-stroke depression over traditional predictors such as age, sex years of education and physical independence (Hackett & Anderson; Barker-Collo, 2007) and can more than double depression risk at 6 months post-stroke (Williams & Demeyere, 2021). Though post-stroke anxiety is less frequently researched, some studies report associations to objective cognitive impairment (Fure et al., 2006; Shimoda, 1998; Tang et al., 2021).

Post-stroke depression and anxiety are not only related to objective cognitive performance on standardized tests. Subjective cognitive complaints, defined as whether individuals self-report cognitive difficulties, are thought to represent emotional concern about cognitive impairment. In systematic reviews, subjective cognitive complaints have little to no association with objective cognitive performance (van Rijsbergen et al., 2014), but have a stronger correlation with post-stroke depression (Duits et al., 2008; Nijsse et al., 2017), work and leisure activities (McKevitt et al., 2011), self-efficacy (Aben et al., 2011) and fatigue (Maaijwee et al., 2014; van Rijsbergen et al., 2018), all of which impact quality of life. Subjective cognitive complaints are important because they tend to increase over time post-stroke (van Rijsbergen et al., 2014) and can be a precursor to cognitive decline and dementia risk (Mitchell, 2008; Juncos-Rabadan et al., 2012). Outside of stroke, theoretical models of anxiety (Koerner & Dugas, 2006) and depression (Beck, 1974) state that negative subjective evaluations and interpretations of abilities, including cognitive abilities such as forgetfulness and inattention, increase risk of emotional distress, and over time, worsen depression and anxiety symptoms. Given this, subjective cognitive complaints may be a risk factor for poorer long-term emotional outcomes post-stroke.

Though objective cognitive performance relates to depression cross-sectionally within early stroke (Maaijwee et al., 2014; Williams & Demeyere, 2021; Hackett et al., 2005), subjective cognitive complaints may be a stronger correlate (van Rijsbergen et al., 2014). Comparatively, the relationship between objective and subjective cognition and anxiety has been less frequently researched. In a systematic review, Menlove et al. (2015) reported that objective cognitive impairment is associated with concurrent anxiety post-stroke. However, to our knowledge, a potential link between subjective cognition and post-stroke anxiety has not yet been examined. Importantly, objective and subjective cognition have not been considered together as predictive factors for post-stroke mood. Examining these variables together in understanding post-stroke mood outcomes is important given the common risk of negative subjective evaluations to depression and anxiety severity outside of stroke (Kircanski et al., 2012; Cisler & Koster, 2010), and overlap between depression and anxiety symptoms post-stroke (Barker-Collo, 2007; Wright et al., 2017).

Finally, links between objective and subjective cognition to post-stroke mood have predominantly been examined cross-sectionally. Existing longitudinal research has focused only on objective cognitive impairments (Nys et al., 2006; Downhill & Robinson, 1994) and within the first-year post-stroke (Lo et al., 2022). Longitudinal data allows for modelling within-individual change over time, rather than comparing potentially very different stroke cohorts cross-sectionally at varying time points, and can provide stronger evidence for potential causality. Importantly, the documented improvements in objective cognitive abilities, and changes in subjective cognitive complaints over time post-stroke (Hochestenbach et al., 2003; van Rijsbergen et al., 2014) may impact depression and anxiety outcomes.

## Study Aim

The study aim was to determine whether changes in subjective cognitive complaints and objective cognitive impairment from 6 months to long-term (>2 years post-stroke) differentially predict changes in depression and anxiety severity post-stroke.

## Methods

Data collection at 6 months (Oxford Cognitive Screening Programme; Reference 14/LO/0648 & 18/SC/0550) and in long-term stroke (OX-CHRONIC Study; Reference 19/SC/0520) was approved by National Research Ethics Committees. All participants provided informed consent to take part.

### Participants and Data Structure

This study is a secondary observational analysis of the OX-CHRONIC study, a longitudinal stroke cohort of post-stroke psychological outcomes (Demeyere et al., 2021; Kusec et al 2023). Participants were initially recruited from the John Radcliffe Hospital acute stroke ward in Oxford, UK as part of the Oxford Cognitive Screening programme (2012 – 2020). Participants consenting to future research opportunities at a 6-month follow-up assessment (Time 1) and who were ≥2 years post-stroke (*N* = 208) were approached for participation in OX-CHRONIC. Participants consenting to OX-CHRONIC completed a battery of self-report and neuropsychological measures remotely across two time points, one year apart (Time 2 and Time 3). OX-CHRONC data were combined with 6-month assessment data to form the current dataset. Participant demographic information and stroke characteristics were obtained from hospital records with participant consent.

### Study Measures

Participants completed the below measures at each time point.

Depression and anxiety were assessed using the *Hospital Anxiety and Depression Scale* (HADS; Zigmond & Snaith, 1983), a 14-item measure (7 items HADS-Depression; 7 items HADS-Anxiety). Each item is rated on 0 – 3 Likert scale. Higher scores indicate greater symptom severity.

Subjective cognitive complaints were assessed using the *Cognitive Failures Questionnaire* (CFQ; Broadbent et al., 1982), a 25-item measure of day-to-day cognitive difficulties (e.g., in memory, attention, perception). Items are answered on a 0 – 4 Likert scale. Higher scores indicate greater subjective cognitive complaints.

Objective cognitive impairment was assessed using the *Oxford Cognitive Screen* (OCS; Demeyere et al., 2015), a validated stroke-specific measure of cognition in domains of memory, spatial attention, praxis, numeracy, language, and executive function. Long-term follow up at Time 2 and 3 used the Tele-OCS (Webb et al 2023), which contains 10 OCS subtests (picture naming, semantics, sentence reading, orientation, verbal memory, episodic memory, number writing, calculations, hearts cancellation, and trail making). Participants scoring < 5th centile relative to normative performance by healthy adults were considered impaired in that subtest. Total proportion of subtests impaired was used as a measure of cognitive impairment severity (e.g., Milosevich et al 2023).

### Data Management and Sample Power

For any self-report questionnaire with < 25% missing item-level data, totals were estimated using mean imputation based on responses from non-missing items. Any participant with ≥25% of HADS or CFQ items missing was treated as a missing observation for that questionnaire. Proportion of OCS subtests impaired was used in analyses (versus number of subtests impaired) to accommodate for any missingness. CFQ scores and OCS proportion of subtests impaired were standardized using z-scores to allow for direct comparisons between independent predictors in analyses.

In a post-hoc power analysis, the sample was adequately powered to detect a moderate effect size of at least 0.31 with 80% power and an alpha of 0.05.

### Statistical Analyses

Analyses were performed using R version 4.2.1 (R Core Team, 2022). Analysis code can be found at https://osf.io/6hdcs/. OX-CHRONIC study data has been deposited in full to Dementia Platforms UK where it can be accessed: https://portal.dementiasplatform.uk/

Descriptive statistics for demographic variables were calculated. Depression and anxiety prevalence rates from 6-months to long-term stroke are reported. To characterize depression and anxiety trajectories from 6 months to long-term stroke, and the degree to which they were predicted by post-stroke demographic variables, “baseline” mixed-effects models were conducted using the *nlme* R package (Pinheiro et al., 2022). We then conducted the final longitudinal mixed-effects models to determine whether changes in objective and subjective cognition over time predicted depression and anxiety severity above the baseline model.

Because depression and anxiety severity, as well as subjective cognitive complaints, are assessed by self-report, it is possible that these measures may be (at least in part) reflecting the same underlying construct. To determine the direction of influence between these measures, exploratory longitudinal mixed-effects models were conducted in reverse (depression and anxiety predicting subjective cognitive complaints) and examined relative to models conducted above.

For all mixed-effects models, a random intercepts, random slopes structure (with participant ID grouped by time point) was used. Using the *performance* R package (Ludecke et al., 2021), adjusted intraclass correlation coefficients (ICCs) were estimated to determine random effects variance (Nakagawa et al., 2017), and marginal R^2^ for fixed effects variance. Model fit statistics including the Akaike Information Criterion (AIC), the Bayesian Information Criterion (BIC), the Root Mean Squared Error (RMSE) and the Sigma (Σ) are reported, with lower values in each statistic indicating better fit. Mixed-effects models are robust to missing datapoints and provide unbiased model parameter estimates with all available information using maximum likelihood estimation. Therefore, all individuals who contributed data were included in analyses, except those lost to attrition due to death as this was not considered to meet the Missing at Random assumption of mixed-effects models.

In addition to mixed effects models, cross-sectional correlations between objective and subjective cognition to depression and anxiety severity, age, time post-stroke, stroke severity, years of education, and participant sex were conducted to aid interpretation and for full transparency of the data.

## Results

*N =* 105 stroke participants were recruited to OX-CHRONIC (see Kusec et al., 2023, for a detailed description), with 6 lost to attrition due to death^1^ (present study *N* = 99; Table 1). Participants predominantly had ischaemic strokes (82.8%) and moderate stroke severity (median NIHSS = 5, Interquartile Range = 6.75).

**Table 1.** Participant Characteristics. Scores higher than 7 on the HADS-D and HADS-A were considered clinically elevated (i.e., warranting clinical attention), as per published cutoffs. HADS = Hospital Anxiety and Depression Scale-Depression; NIHSS = National Institute of Health Stroke Severity

| Participants ($N = 99$) | | Min - Max | % Clinically Elevated |
| --- | --- | --- | --- |
| **Sex – $n$ (%)** |  |  |  |
| Male | 56 (56.6%) |  |  |
| Female | 43 (43.4%) |  |  |
| **Age at Stroke – Mean ($SD$)** | 68.9 (13.1) | 18 – 92 |  |
| **Years of Education – Mean ($SD$)** | 13.9 (3.7) | 9 – 23 |  |
| **Stroke Type – $n$ (%)** |  |  |  |
| Ischaemic | 82 (82.8%) |  |  |
| Haemorrhagic | 17 (17.2%) |  |  |
| **Lesion Hemisphere – $n$ (%)** |  |  |  |
| Left | 40 (40.4%) |  |  |
| Right | 39 (39.4%) |  |  |
| Bilateral | 7 (7.1%) |  |  |
| Undetermined from scan | 13 (13.1%) |  |  |
| **First or Recurrent Stroke – $n$ (%)** |  |  |  |
| First | 67 (67.7%) |  |  |
| Recurrent | 32 (32.3%) |  |  |
| **Acute NIHSS Score – Mean ($SD$)** | 7.5 (6.4) | 0 – 27 |  |
| **HADS-Depression – Mean ($SD$)** |  |  |  |
| Time 1 (6 Months post-stroke) | 4.7 (3.5) | 0 – 16 | 20.5% |
| Time 2 (~4.5 years post-stroke) | 4.7 (3.6) | 0 – 17 | 23.4% |
| Time 3 (~5.5 years post-stroke) | 4.8 (3.6) | 0 – 18 | 18.2% |
| **HADS-Anxiety – Mean ($SD$)** |  |  |  |
| Time 1 (6 Months post-stroke) | 5.3 (3.6) | 0 – 16 | 24.5% |
| Time 2 (~4.5 years post-stroke) | 5.1 (3.6) | 0 – 15 | 22.4% |
| Time 3 (~5.5 years post-stroke) | 5.3 (3.9) | 0 – 20 | 26.1% |

Cross-sectional correlations between HADS subscales to OCS and CFQ scores are in Figure 1. At all time points, objective cognition and subjective cognitive complaints did not correlate with each other, nor with age, time-post stroke or years of education (*p*s = 0.13 – 0.94) nor were there any observed sex differences (see *Supplementary Materials* for details).

**Figure 1.**
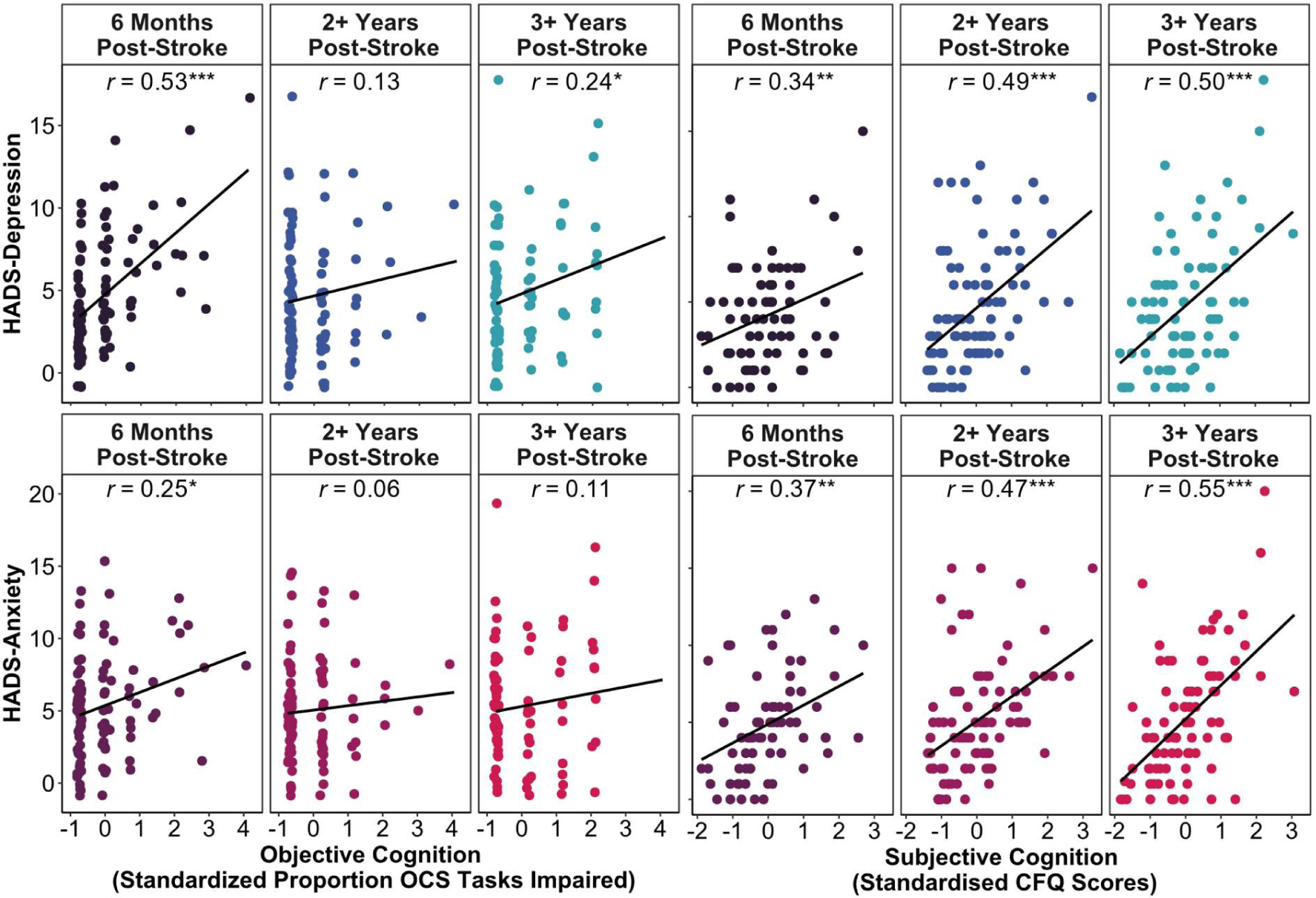
Cross-sectional pairwise Pearson correlations between objective and subjective cognition to depression severity (top row) and to anxiety severity (bottom row) across all study time points. OCS = Oxford Cognitive Screen; CFQ = Cognitive Failures Questionnaire; HADS = Hospital Anxiety and Depression Scale * *p* < 0.05, ** *p* < 0.01, *** *p* < 0.001

### Baseline Model – Depression and Anxiety Trajectories

There was no mean change in post-stroke depression from Time 1 to Time 2 (*b* = - 0.04, SE = 0.33, *p* = .90; VC = 7.91, *SD* = 2.81) nor to Time 3 (*b* = 0.24, SE = 0.36, *p* = .51; VC = 9.34, *SD* = 3.06; ICC_adj_ = .89 [AIC = 1396.93, BIC = 1433.17, marginal R2 = 0.001, RMSE = 0.59, Σ = 1.21]). Similarly, there was no change in anxiety scores to Time 2 (*b* = - 0.35, SE = 0.38, *p* = .36; VC = 9.89, *SD* = 3.15) or Time 3 (*b* = -0.02, SE = 0.39, *p* = .0.96; VC = 10.39, *SD* = 3.22; ICC_adj_ = .87 [AIC = 1442.54, BIC = 1478.78, marginal R2 = 0.002, RMSE = 0.64, Σ = 1.34]). Depression and anxiety were thus stable on average from 6 months to chronic stroke. However, there appeared to be large variation in both improvement and worsening subgroups between timepoints (Figure 2).

**Figure 2.**
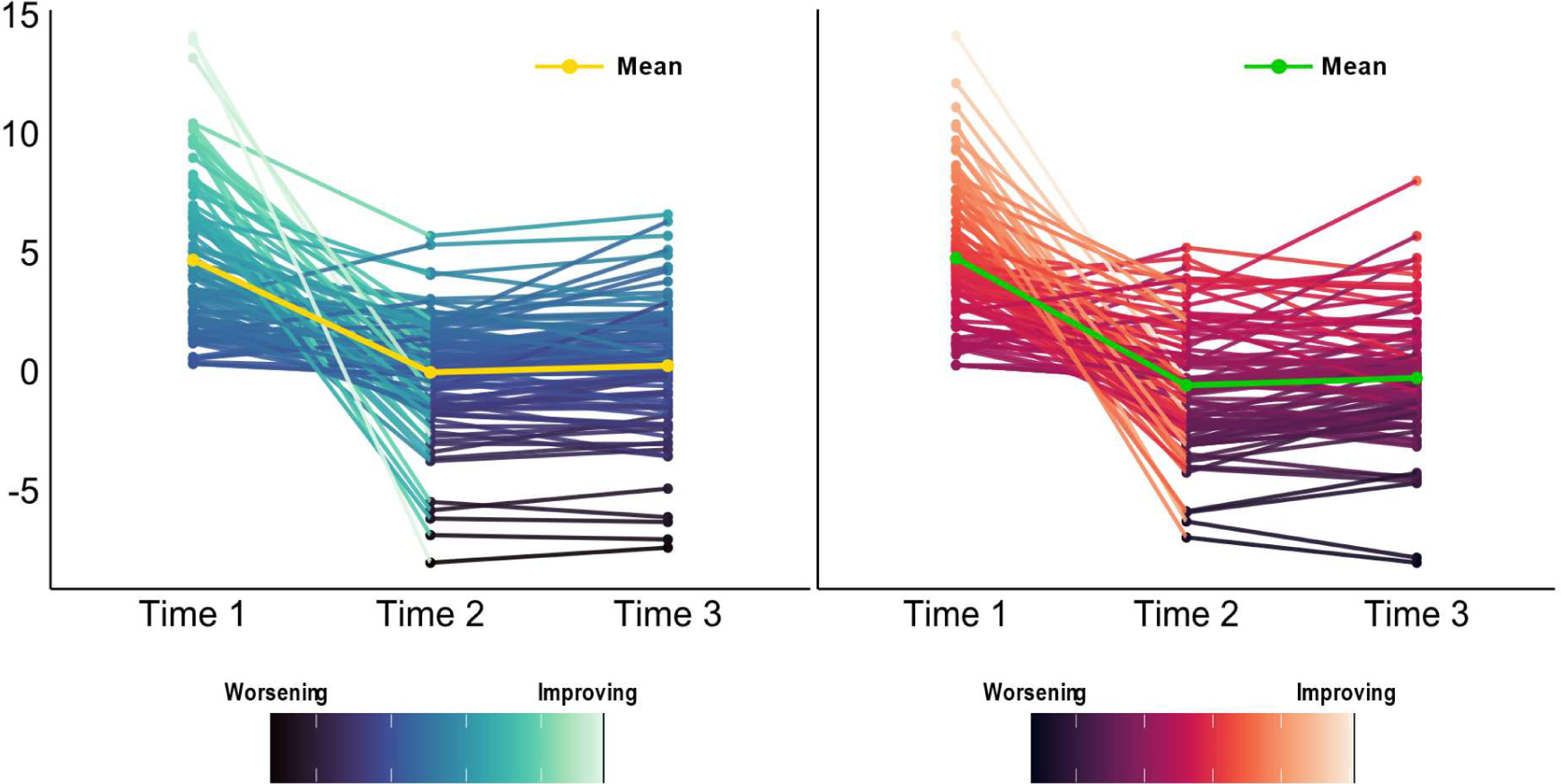
Participant-level trajectories of depression (left hand panel) and anxiety severity (right hand panel) from 6-months (Time 1) to approximately 4.5 (Time 2) and 5.5 years (Time 3) post-stroke. The y-axis shows individual-level change in HADS-D and HADS-A after accounting for expected change over time and between-participant variability (random slopes). Darker colours represent worse depression or anxiety and lighter colours represent better scores. Mean trajectory values are shown in yellow for depression severity, and in green for anxiety severity.

When adding demographic variables to baseline models, HADS-Depression and HADS-Anxiety scores were not predicted by acute NIHSS scores (HADS-D *b* = 0.08, SE = 0.06, *p =* .19; HADS-A *b* = 0.05, SE = 0.05, *p =* .34), participant concurrent age (HADS-D *b* = 0.01, SE *=* 0.03, *p* = 0.79; HADS-A *b* = -0.03, SE = 0.03, *p =* .26), years of education (HADS-D *b* = -0.10, SE = 0.09, *p =* .30; HADS-A *b* = -0.18, SE = 0.09, *p =* .07), or participant sex (HADS-D *b* = -0.30, SE = 0.72, *p =* .67; HADS-A *b* = -0.17, SE = 0.71, *p =* .81). Inclusion of demographic variables explained little variance and did not improve fit for HADS-Depression (marginal R^2^ = 0.03; AIC = 1150.84, BIC = 1198.54; RMSE = 0.60, Σ = 1.24; ICC_adj_ = 0.88) nor HADS-Anxiety (marginal R^2^ = 0.05; AIC = 1196.93, BIC = 1244.63; RMSE = 0.65, Σ = 1.39; ICC_adj_ = 0.88).

### Longitudinal Association of Objective and Subjective Cognition to Depression

When including objective and subjective cognition to the baseline depression model, both objective (*B* = 1.01, SE = 0.29, *p* < .001) and subjective cognition (*B* = 0.83, SE = 0.27, *p* < .01) had main effects on depression severity, indicating a stable relationship between these variables regardless of timepoint.

In terms of interaction effects, objective cognitive impairments significantly related to HADS-Depression scores at 6-months but this weakened at 4.5 years (*B* = -0.79, SE = 0.37, *p* < .05). The impact of subjective cognition to HADS-Depression did not change between Time 1 to Time 2, though a trend was noticed (*B* = 0.60, SE = 0.34, *p* = .08). Subjective cognitive complaints at Time 3 appeared to have a stronger effect on HADS-D scores (*B* = 0.77, SE = 0.35, *p* < .05). Inclusion of the interaction term between subjective cognitive complaints over time provided the best fit to the study data (see Figure 2; AIC = 1160.15, BIC = 1215.90, RMSE = 0.43, Σ = 0.99, marginal R^2^ = 0.22) compared to modelling only an interaction between objective cognition over time (AIC = 1334.99, BIC = 1381.73, RMSE = 0.52, Σ = 1.12, marginal R^2^ = 0.08). Variance due to random slopes was moderate at Time 2 (VC = 5.04, *SD* = 2.25) and Time 3 (VC = 6.29, *SD* = 2.51), with an ICC_adj_ of 0.89.

Exploratory analyses were conducted investigating whether changes in any specific cognitive domains predicted depression (see *Supplementary Materials* for full results). Only attention abilities related to depression outcomes at Time 1 relative to Time 2 (*b* = -1.81, SE = 0.81, *p* < .05) and Time 3 (*b* = -1.90, SE = 0.80, *p* < .05).

### Longitudinal Association of Objective and Subjective Cognition to Anxiety

Only subjective cognition had a main effect on HADS-Anxiety scores (*B* = 1.14, SE = 0.34, *p* < .01) whereas objective cognition did not (*B* = 0.32, SE = 0.37, *p* = .40).

This lack of relationship between objective cognition and HADS-Anxiety remained constant from Time 1 to Time 2 (*B* = -0.49, SE = 0.48, *p* = .30) and from Time 2 to Time 3 (*B* = -0.15, SE = 0.45, *p* = .73). Though the relationship between subjective cognition and HADS-Anxiety from Time 1 to Time 2 was stable (*B* = 0.17, SE = 0.44, *p* = .69), this relationship strengthened at Time 3 (*B* = 0.96, SE = 0.44, *p* < .05). Variance due to random slopes was moderate at Time 2 (VC = 10.62, *SD* = 3.26) and Time 3 (VC = 9.76, *SD* = 3.12), with an ICC_adj_ of 0.86. As in the depression model, inclusion of the interaction term between subjective cognitive complaints over time provided the best fit to the data (AIC = 1226.20, BIC = 1281.96, RMSE = 0.52, Σ = 1.19, marginal R^2^ = 0.20; see Figure 2) compared to modelling only an interaction between objective cognition over time (AIC = 1394.48, BIC = 1441.21, RMSE = 0.59, Σ = 1.23, marginal R^2^ = 0.02).

Exploratory domain-specific cognitive impairment models were not conducted due to objective cognitive impairment not demonstrating a main effect in anxiety severity.

### Exploring Direction of Influence Between Mood and Subjective Cognition

It is possible that a relationship between subjective cognition to depression and anxiety exists because self-report measures capture similar response tendencies. Cognitive theories of depression and anxiety (Beck, 1974, Koerner & Dugas, 2006) state that overly negative evaluations of abilities increases negative affect, instead of the reciprocal. When conducting models in reverse, there were no interaction between changes in HADS-Depression from Time 1 to Time 2 (*B* = 0.05, SE = 0.12, *p* = .70) or at Time 3 (*B* = -0.01, SE = 0.12, *p* = .94) to changes in subjective cognition. Similarly, there were no interaction effects in changes in HADS-Anxiety over time from Time 1 to Time 2 (*B* = -0.03, SE = 0.03, *p* = 0.33) nor at Time 3 (*B* = -0.05, SE = 0.03, *p* = 0.13). See the *Supplementary Materials* for a full account.

### Summary of Results

Taken together, objective cognitive impairments, in particular attention impairments, at 6 months post-stroke were related to depressive symptoms only. However, this relationship weakened in long-term stroke (∼4.5 years). Subjective cognitive complaints had a stronger relationship to both depression and anxiety in long-term (≥2 years) stroke compared to 6-months post-stroke (Table 2). When exploring potential direction of influence between subjective cognitive complaints to mood outcomes, changes in depression and anxiety severity did not have a reciprocal effect on subjective cognition.

**Table 2.** Mixed-effects model results. Bolded text indicates statistically significant terms. OCS: Oxford Cognitive Screen; CFQ = Cognitive Failures Questionnaire

| Variable | HADS-Depression |  | HADS-Anxiety |  |
| --- | --- | --- | --- | --- |
|  | *B* (SE) | *t* (*p*-value) | *B* (SE) | *t* (*p*-value) |
| Intercept | **4.26 (0.30)** | **14.14 (<0.001)** | **4.99 (0.35)** | **14.15 (<0.001)** |
| Time 2 | 0.34 (0.31) | 1.11 (0.27) | -0.03 (0.42) | -0.07 (0.94) |
| Time 3 | 0.57 (0.34) | 1.69 (0.09) | 0.32 (0.42) | 0.76 (0.45) |
| Proportion OCS Tasks Impaired | **1.01 (0.29)** | **3.48 (<0.001)** | 0.32 (0.37) | 0.85 (0.40) |
| CFQ | **0.83 (0.27)** | **3.08 (<0.01)** | **1.14 (0.34)** | **3.34 (<0.01)** |
| Time 2 * Proportion OCS Tasks Impaired | **-0.79 (0.37)** | **-2.13 (<0.05)** | -0.49 (0.48) | -1.03 (0.30) |
| Time 3 * Proportion OCS Tasks Impaired | -0.43 (0.36) | -1.19 (0.24) | -0.16 (0.45) | -0.35 (0.73) |
| Time 2 * CFQ | 0.60 (0.34) | 1.79 (0.08) | 0.17 (0.44) | 0.40 (0.69) |
| Time 3 * CFQ | **0.77 (0.35)** | **2.19 (<0.05)** | **0.96 (0.43)** | **2.17 (<0.05)** |

**Figure 3.**
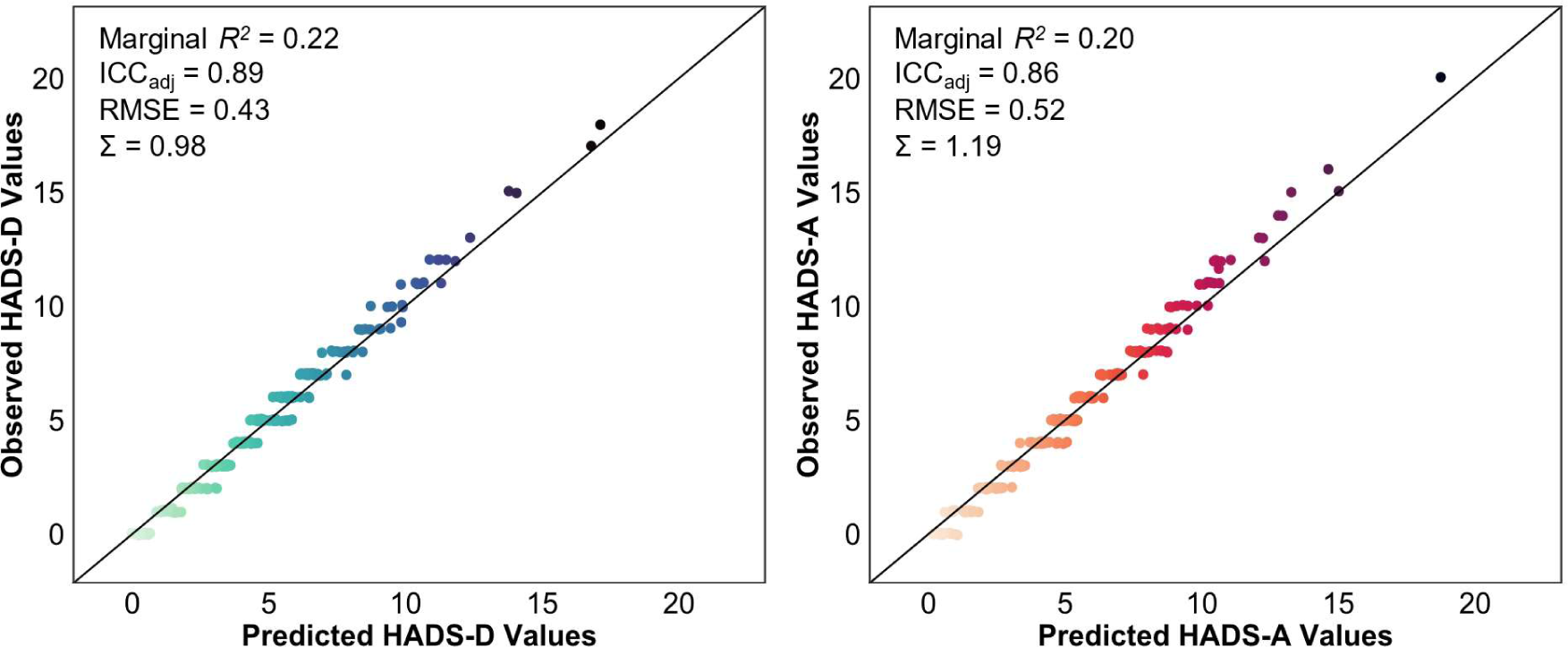
Predicted versus observed values of HADS-D scores (left) and HADS-A scores (right). Predicted values are based on model fit indices. HADS-D = Hospital Anxiety and Depression Scale – Depression; HADS-A = Hospital Anxiety and Depression Scale – Anxiety; ICC = Intraclass Correlation Coefficient; RMSE = Root Mean Square Error

## Discussion

This study examined whether the relationship between objective cognitive impairment and subjective cognitive complaints to post-stroke depression and anxiety differed from 6 months to long-term stroke (average ∼5.5 years, range 2 – 10 years). We found that typical predictors of post-stroke function such as age, years of education, sex, and stroke severity did not relate to post-stroke depression and anxiety (collective marginal R^2^ = 0.03 – 0.05). Instead, depression and anxiety severity was better explained by objective and subjective cognition (marginal R^2^ = 0.20 – 0.22). While both objective and subjective cognition related to post-stroke depression, only subjective cognition related to post-stroke anxiety. Specifically, objective cognitive impairments at 6 months post-stroke related to depression, but this effect weakened in long-term stroke. Subjective cognitive complaints mostly strongly influenced post-stroke depression and anxiety in long-term stroke.

### Relationship between Objective and Subjective Cognition Over Time

Though both objective cognitive impairment and subjective cognitive complaints related to depression and anxiety severity, they did not relate to each other at any time point. Many studies report no correlation between objective cognitive impairments and subjective cognitive complaints both in acute stroke (Kliem et al., 2022; Duits et al., 2008) and onwards post-stroke (Lamb et al., 2013). This may be explained by the content of subjective cognition self-report measures. In systematic reviews, the highest correlation between subjective cognitive complaints and objective performance is on more salient cognitive impairments such as memory (Davis et al., 1995; Duits et al., 2008), whereas executive function difficulties for example may be more difficult to self-report. However, outside of stroke, meta-analytic evidence suggests only a modest correlation between self-reported abilities and objective performance (*r* = 0.29, Zell & Krizan, 2014), indicating that discrepancies may be due to pre-stroke difficulties in self-assessing abilities.

The weak relationship between objective and subjective cognition may indicate that each has distinct influences on post-stroke depression and anxiety. Our findings on objective cognitive impairment align with previous research showing that improvements in cognitive ability enhances function and thus mood (Mole & Demeyere, 2020; Wheeler et al., 2023). It is less clear how subjective cognition changes affect mood outcomes. Lowered access to psychological care in the long-term post-stroke (Lin et al., 2021) may result in increased distress over unaddressed subjective cognitive complaints. Alternatively, self-reports of cognitive difficulties may represent worry about possible cognitive decline post-stroke (Tang et al., 2019). Qualitative literature has highlighted that uncertainty about future cognitive status can increase post-stroke anxiety (Salter et al., 2008). These increases in uncertainty in cognitive abilities may explain the stronger relationship between subjective cognitive complaints and mood outcomes in longer-term stroke.

### Objective Cognitive Performance and Post-Stroke Depression and Anxiety

We found that the relationship between objective cognitive impairment to post-stroke depression severity is strongest at 6 months post-stroke and lessens into the long term. The relationship between objective cognitive impairments and post-stroke depression is well-documented in systematic reviews (Hackett & Anderson, 2005; Ayerbe et al., 2013; Chun et al., 2022). Objective cognitive impairments are more prevalent at 6 months compared to long-term stroke (Kusec et al., 2023), therefore recovery made may have a more noticeable influence and therefore improve depression, relative to potentially minor shifts in objective cognitive ability in long-term stroke. However, there are alternative explanations. First, although cognitive impairment is still common in long-term stroke (∼45% prevalence; Kusec et al., 2023), it may no longer be severe enough to substantially impact self-rated depression. Second, individuals may become more adept at managing the impact of cognitive impairment on daily life (Ch’ng et al., 2008), weakening its link to depression. Finally, improvements in specific cognitive impairments may better explain improvements in mood status than overall severity of cognitive impairment, as evidenced in cross-sectional research (Williams & Demeyere, 2021). In our exploratory analyses, we found that only attention impairments relate to depression outcomes from 6 months to long-term stroke. Though replication is warranted, this result aligns with research outside of stroke, where concentration difficulties and increased distractibility are core features of depression (Keller et al., 2019). Possibly, attentional impairments aggravate depression-related concentration and distractibility symptoms and worsen depression, though further explorations of this in post-stroke depression are warranted.

We found no role of objective cognitive impairment to post-stroke anxiety. The relationship between these two variables has been inconsistent, with some research demonstrating no unique correlation between these variables (Scopelliti et al., 2022; Williams & Demeyere, 2021) and others reporting strong predictive effects of objective cognitive performance to post-stroke anxiety in both early (Barker-Collo, 2007) and long-term stroke (Lee et al., 2019). Given their inconsistent relationship, larger scale research is needed prior to accepting or rejecting a cognition-anxiety relationship. Additionally, we found that demographic factors do not reliably predict post-stroke anxiety, contrary to previous research demonstrating younger age as a risk factor (Menlove et al., 2015). Possibly, demographic factors contribute to only a small amount of variance in anxiety (as found here) that are only statistically detectable in meta-analyses with far larger samples (e.g., Chun et al., 2022).

### Subjective Cognitive Complaints and Post-Stroke Depression and Anxiety

Subjective cognitive complaints have long been associated with post-stroke depressive symptoms (van Rijsbergen et al., 2014). Possibly, in the very long-term post-stroke, individuals become more adept at *recognizing* cognitive changes, and begin ruminating on the implications of this on everyday life. Alternatively, long-term stroke may result is greater social isolation due to decreases in occupational and leisure activities (Mole & Demeyere, 2020; Stolwyk et al., 2021). Greater social isolation can result in more time ruminating on stroke-related changes, including increased focus on cognitive changes (White et al., 2008; Turner et al., 2019).

We found that *only* changes in subjective cognitive complaints related to post-stroke anxiety, particularly in long-term stroke. Similar to the depression models, changes in anxiety post-stroke did not reciprocally relate to changes in subjective cognitive complaints. Outside of post-stroke depression, the role of subjective cognitive complaints has only been investigated in post-traumatic stress disorder symptoms (Menlove et al., 2015). In qualitative research, stroke survivors report difficulties accepting not being able to predict if and how their cognitive abilities may change in the future, leading to worry (Carlsson et al., 2009; Crowe et al., 2016). Further quantitative research has shown that difficulties coping with uncertainty are uniquely associated with post-stroke anxiety (Kusec et al., 2023) as has been shown outside of stroke (Koerner & Dugas, 2006). Identifying mediators of the relationship between subjective cognitive complaints and post-stroke anxiety can be a fruitful avenue for research.

Elevated reports of subjective cognitive complaints could also represent an “over-awareness” of post-stroke abilities – that is, endorsing a relatively mild cognitive change as distressing despite objective performance corresponding to expected, or pre-stroke, performance (e.g., Wheeler et al., 2023; Smeets et al., 2017). Another explanation is that *pre-stroke* negative evaluation tendencies extend to post-stroke abilities more generally (Beck, 1974; Koerner & Dugas, 2006). Whether negative evaluations are unique to cognitive difficulties post-stroke, compared to physical abilities for example, would help determine whether a more general versus cognition-specific negative evaluation tendency is important to depression and anxiety outcomes post-stroke.

Subjective cognitive complaints are thought to be manifestations of depressed mood (Nijsse et al., 2017) rather than independent from depression. Notably, we found that whilst depression and anxiety severity are correlated with subjective cognition over time, changes in depression and anxiety severity did not reciprocally relate to changes in subjective cognition. These findings are in line with cognitive models of depression in non-stroke populations, in which negative evaluations of abilities worsen depression outcomes (Beck, 1974) rather than the reverse. Possibly, subjective cognitive complaints have a more context-dependent or dynamic relationship to depression and anxiety, while the effect of depression/anxiety on subjective cognition is comparably constant over time.

## Clinical Implications

A top unmet need of long-term stroke survivors is mental health support (Lin et al., 2021; Hill et al., 2022). Using longitudinal analyses, we have demonstrated addressing subjective or objective cognition in mood interventions may depend on time-point post-stroke. At 6 months post-stroke, including cognitive rehabilitation strategies within mood interventions, specifically attentional management strategies, may be beneficial for depression. However, in long-term (>2 years) stroke, managing subjective cognitive complaints may be more valuable to incorporate into both depression and anxiety interventions. Our results hold particular promise for developing anxiety interventions, as subjective cognitive complaints are potentially modifiable, unlike other reported risk factors such as age and sex (Chun et al., 2022).

In developing mood interventions for long-term stroke, subjective cognitive complaints likely relate to depression and anxiety severity differently. Depression is characterized by ruminative tendencies over past events, while anxiety is characterized by pathological worry about events that may occur in the future. Individuals with post-stroke depression may ruminate over the loss of cognitive function compared to pre-stroke abilities, while those with post-stroke anxiety worry about whether cognitive abilities will change or deteriorate in the future. These distinctions may result in similar scores on subjective cognitive complaints measures, but could have different treatment implications (i.e., in targeting the content of concern driving such reporting).

## Limitations

Though time post-stroke did not correlate with changes in depression and anxiety, data time points 2 and 3 comprised a wide range of participants from 2 to 9 years post-stroke. Although pre-stroke emotional status is known to relate to post-stroke mood outcomes, such data was not available here. Finally, the OCS and the HADS are meant to be screening tools rather than diagnostic instruments, thus longitudinal results should be replicated with repeated assessments of neuropsychological batteries and interview-based mood disorder diagnoses.

## Conclusions

Objective cognitive impairment and subjective cognitive complaints better predict changes in depression symptoms from 6 months to the long-term post-stroke than demographic variables. Subjective cognitive complaints relate uniquely to post-stroke anxiety. This research may have clinical implications regarding (i) the inclusion of cognitive strategies for depression interventions at 6 months post-stroke and (ii) addressing subjective cognitive complaints in both depression and anxiety interventions in long-term stroke.

## Data Availability

Study analysis code can be found at https://osf.io/6hdcs/. OX-CHRONIC study data has been deposited in full to Dementia Platforms UK where it can be accessed: https://portal.dementiasplatform.uk/

## Acknowledgements

The authors would like to acknowledge Elise Milosevich, Owen A. Williams, Evangeline G.Chiu, Pippa Watson, Bogna A. Drozdowska, Avril Dillon, Trevor Jennings, Bloo Anderson, Helen Dawes, Shirley Thomas, Annapoorna Kuppuswamy, Sarah T. Pendlebury, and Terence J. Quinn for their work as part of the OX-CHRONIC group.

## Ethical Standards

The authors assert that all procedures contributing to this work comply with the ethical standards of the relevant national and institutional committees on human experimentation and with the Helsinki Declaration of 1975, as revised in 2008.

## Footnotes

1 There were no differences in demographics and study measures between those lost to attrition due to death by Time 3 (*N* = 6) and those retained (*N* = 99) (see *Supplementary Table 1*).

## Supplementary Materials

*Differences in Time 1 (6-months post-stroke) between those Retained and those Lost to Attrition Due to Death*

**Supplementary Table 1.** Mean and frequency statistics of demographic, cognitive, and stroke outcome data for participants that were retained across time points (*N* = 99) and those who withdrew due to death (*N* = 6) at Time 1 (6-months post-stroke). NIHSS = National Institute of Health Stroke Severity; OCS = Oxford Cognitive Screen; CFQ = Cognitive Failures Questionnaire; HADS = Hospital Anxiety and Depression Scale * *p* < 0.05

| Measure | Retained (N = 99) | Died (N = 6) | P-Value |
| --- | --- | --- | --- |
| **Demographics** |  |  |  |
| Age | $M = 69.46$ (13.10) | $M = 67.84$ (12.84) | $p = 0.77$ |
| Sex | 56.57% Male | 100.00% Male | $p = 0.08$ |
| Stroke Type | 82.82% Ischaemic | 100.0% Ischaemic | $p = 0.59$ |
| Years of Education | $M = 13.93$ (3.65) | $M = 14.17$ (4.36) | $p = 0.88$ |
| Acute NIHSS Score | $M = 7.49$ (6.35) | $M = 5.80$ (4.55) | $p = 0.56$ |
| Time Post-Stroke (Time 2) | $M = 4.48$ (2.10) | $M = 5.91$ (2.31) | $p = 0.11$ |
| **Study Measures** |  |  |  |
| OCS Proportion Tasks Impaired | $M = 0.10$ (0.14) | $M = 0.12$ (0.08) | $p = 0.59$ |
| CFQ Total | $M = 30.52$ (15.96) | $M = 44.19$ (11.14) | $p = 0.15$ |
| HADS-Depression | $M = 4.71$ (3.54) | $M = 8.00$ (5.62) | $p = 0.05$ |
| HADS-Anxiety | $M = 5.32$ (3.55) | $M = 4.67$ (4.50) | $p = 0.67$ |

*Correlations between Objective Cognitive Impairment and Subjective Cognitive Complaints to Stroke Demographic Factors across Time points*

**Supplementary Table 2.** Pairwise cross-sectional correlations between stroke demographic factors and objective and subjective cognition. OCS = Oxford Cognitive Screen; CFQ = Cognitive Failures Questionnaire; NIHSS = National Institute of Health Stroke Severity. * *p* < 0.05

|  | Objective Cognition<br>(OCS Proportion Tasks Impaired) |  |  | Subjective Cognition<br>(CFQ) |  |  |
| --- | --- | --- | --- | --- | --- | --- |
|  | Time 1<br>(6 Months) | Time 2<br>(~4.5 Years) | Time 3<br>(~5.5. Years) | Time 1<br>(6 Months) | Time 2<br>(~4.5 Years) | Time 3<br>(~5.5. Years) |
| **NIHSS** | 0.26* | -0.06 | 0.08 | -0.08 | 0.04 | -0.01 |
| **Age** | -0.09 | 0.02 | 0.08 | -0.01 | -0.07 | 0.01 |
| **Time Post-Stroke** | -- | 0.17 | -0.08 | -- | 0.15 | 0.09 |
| **Years of Education** | -0.12 | -0.07 | -0.13 | 0.05 | -0.06 | -0.08 |
| **Objective Cognition** | -- | -- | -- | 0.11 | 0.22 | 0.07 |
| **Subjective Cognition** | 0.11 | 0.22 | 0.07 | -- | -- | -- |

**Supplementary Table 3.**
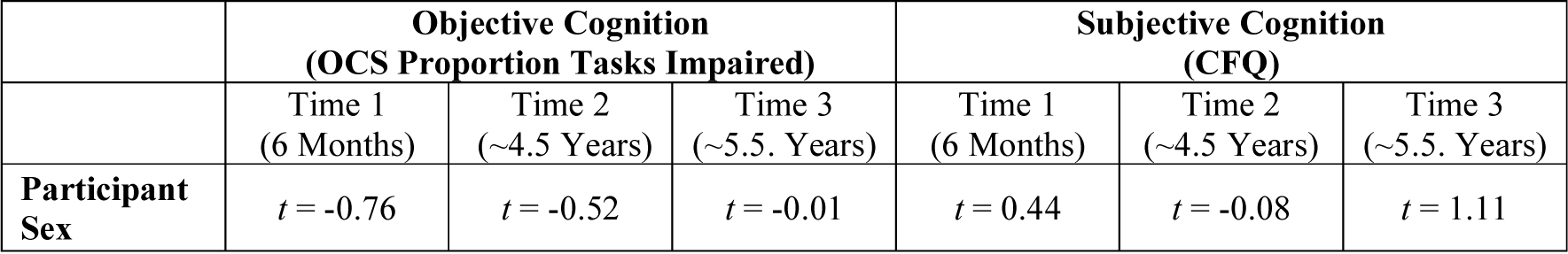
Independent samples t-tests to determine whether objective and subjective cognition differed by participant sex. No t-tests were statistically significant. OCS = Oxford Cognitive Screen; CFQ = Cognitive Failures Questionnaire

*Exploratory Mixed Effects Models of Domain-Specific Impairments Predicting Depression*

**Supplementary Table 4.** Exploratory mixed effects models of the contribution of domain-specific cognitive impairments to depression. Though the interaction effect of changes in subjective cognitive complaints remains across domain-specific models, only changes in attention impairments related to changes in depression severity. * *p* < 0.05, \*\**p* < 0.01, \*\*\**p* <0.001

| HADS-Depression | *b* | *SE* | *t*-value | Pr(>\|*t*\|) |
| --- | --- | --- | --- | --- |
| (Intercept) | 3.94 | 0.35 | 11.37 | <0.001*** |
| Time 2 (~4.5 Years) | 0.58 | 0.34 | 1.71 | 0.08 |
| Time 3 (~5.5 Years) | 0.79 | 0.37 | 2.15 | <0.05* |
| OCS Language Impairment | 1.44 | 0.74 | 1.96 | 0.05 |
| CFQ | 0.86 | 0.28 | 3.03 | <0.01** |
| Time 2*OCS Language Impairment | -0.53 | 1.05 | -0.50 | 0.62 |
| Time 2*OCS Language Impairment | -0.09 | 1.08 | -0.09 | 0.93 |
| Time 2*CFQ | 0.71 | 0.33 | 2.14 | <0.05* |
| Time 3*CFQ | 0.89 | 0.37 | 2.44 | <0.05* |
| (Intercept) | 3.73 | 0.34 | 11.37 | <0.001*** |
| Time 2 (~4.5 Years) | 0.83 | 0.35 | 1.71 | <0.05* |
| Time 3 (~5.5 Years) | 1.13 | 0.39 | 2.15 | <0.01** |
| OCS Attention Impairment | 2.07 | 0.63 | 1.96 | <0.01** |
| CFQ | 0.81 | 0.27 | 3.03 | <0.01** |
| Time 2*OCS Attention Impairment | -1.81 | 0.81 | -0.50 | <0.05* |
| Time 2*OCS Attention Impairment | -1.90 | 0.80 | -0.09 | <0.05* |
| Time 2*CFQ | 0.68 | 0.32 | 2.14 | <0.05* |
| Time 3*CFQ | 0.93 | 0.36 | 2.44 | <0.01** |
| (Intercept) | 4.00 | 0.35 | 11.50 | <0.001*** |
| Time 2 (~4.5 Years) | 0.61 | 0.34 | 1.82 | 0.07 |
| Time 3 (~5.5 Years) | 0.63 | 0.38 | 1.69 | 0.09 |
| OCS Memory Impairment | 1.07 | 0.72 | 1.48 | 0.14 |
| CFQ | 0.80 | 0.29 | 2.79 | <0.01** |
| Time 2*OCS Memory Impairment | -0.94 | 1.11 | -0.85 | 0.39 |
| Time 2*OCS Memory Impairment | 1.35 | 1.05 | 1.28 | 0.20 |
| Time 2*CFQ | 0.69 | 0.34 | 2.03 | <0.05* |
| Time 3*CFQ | 0.99 | 0.36 | 2.74 | <0.01** |

**Supplementary Table 4.** Exploratory mixed effects models of the contribution of domain-specific cognitive impairments to depression.
| **HADS-Depression** | *b* | *SE* | *t*-value | Pr(>\|*t*\|) |
| --- | --- | --- | --- | --- |
| (Intercept) | 4.15 | 0.33 | 12.41 | < **0.001** *** |
| Time 2 (~4.5 Years) | 0.59 | 0.33 | 1.78 | 0.07 |
| Time 3 (~5.5 Years) | 0.75 | 0.37 | 2.02 | < **0.05** * |
| OCS Number Impairment | 0.74 | 1.15 | 0.64 | 0.52 |
| CFQ | 0.87 | 0.29 | 3.02 | < **0.01** ** |
| Time 2*OCS Number Impairment | -1.53 | 1.36 | -1.13 | 0.26 |
| Time 2*OCS Number Impairment | -0.68 | 1.35 | -0.51 | 0.61 |
| Time 2*CFQ | 0.69 | 0.33 | 2.07 | < **0.05** * |
| Time 3*CFQ | 0.92 | 0.37 | 2.54 | < **0.05** * |
| (Intercept) | 3.83 | 0.34 | 11.24 | < **0.001** *** |
| Time 2 (~4.5 Years) | 0.89 | 0.34 | 2.63 | < **0.01** ** |
| Time 3 (~5.5 Years) | 1.18 | 0.37 | 3.23 | < **0.01** ** |
| OCS Executive Impairment | 0.99 | 0.63 | 1.56 | 0.12 |
| CFQ | 0.52 | 0.28 | 1.85 | 0.07 |
| Time 2*OCS Executive Impairment | -1.53 | 0.97 | -1.58 | 0.11 |
| Time 2*OCS Executive Impairment | -2.19 | 1.28 | -1.72 | 0.09 |
| Time 2*CFQ | 1.05 | 0.34 | 3.09 | < **0.01** ** |
| Time 3*CFQ | 1.26 | 0.37 | 3.46 | < **0.001** * |

*Exploratory Models of Depression and Anxiety Predicting Subjective Cognitive Complaints*

**Supplementary Table 5.**
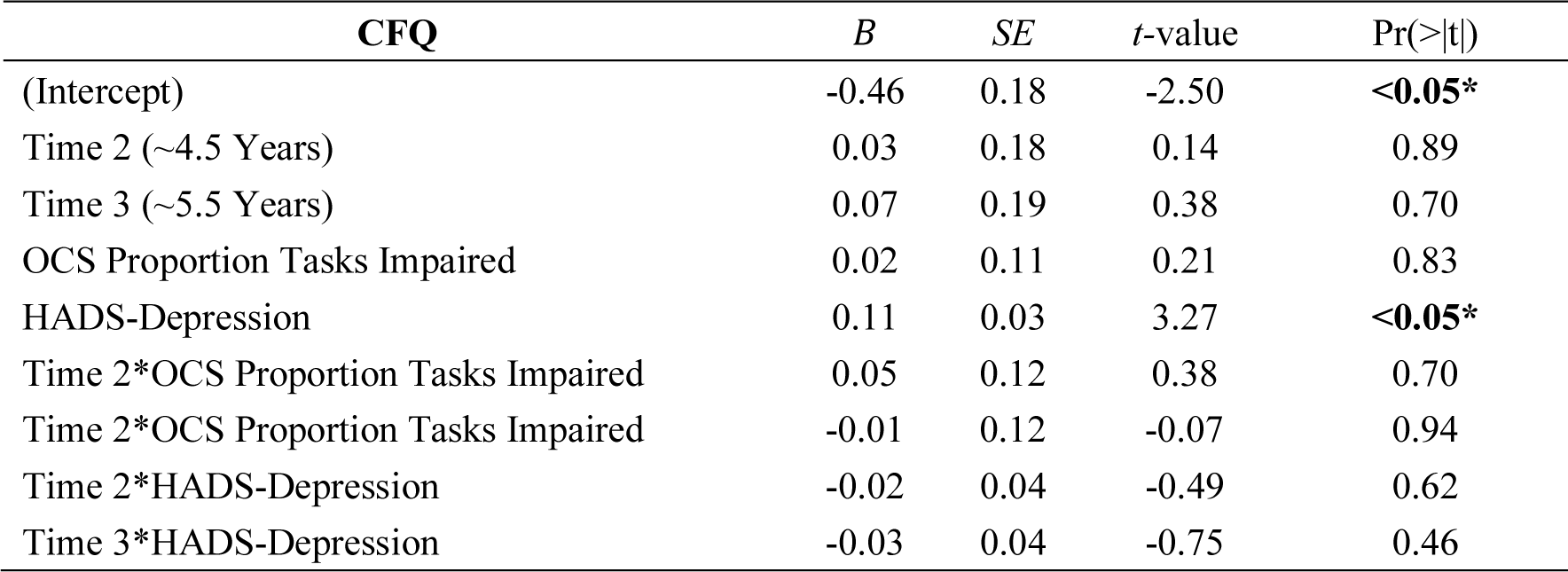
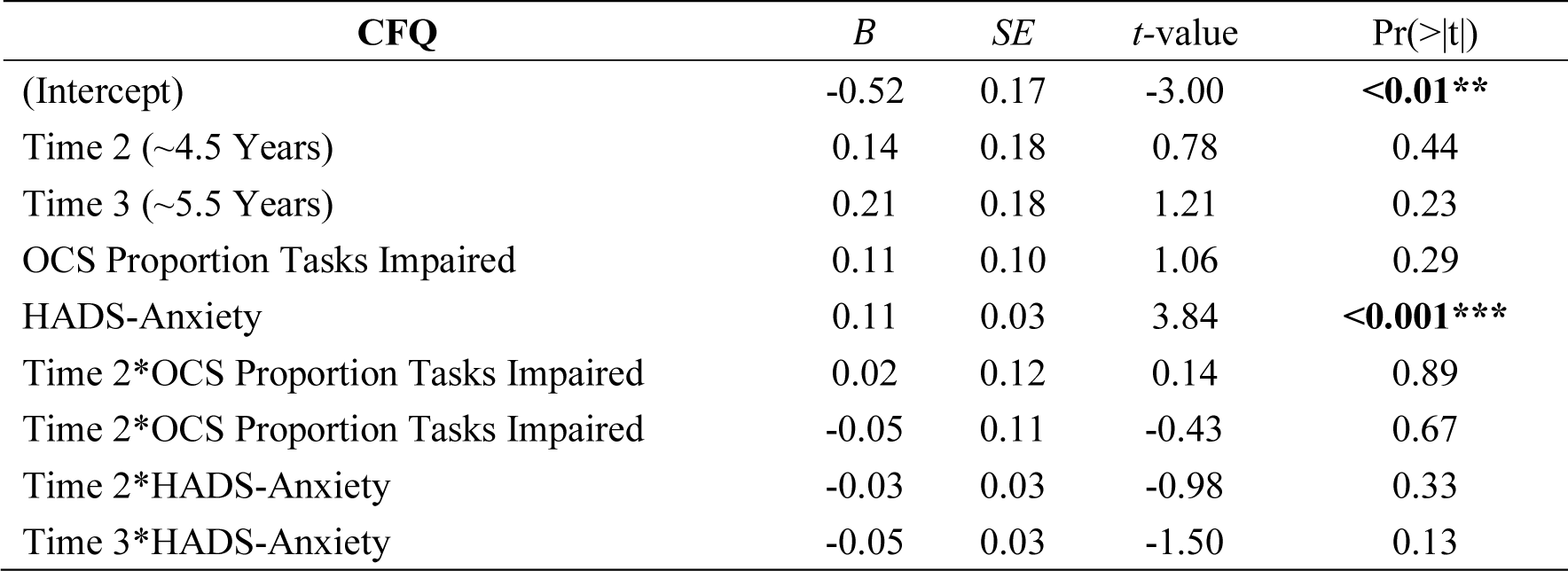
Exploratory mixed effects models with HADS-Depression and HADS-Anxiety scores predicting changes in subjective cognitive complaints. Unlike the main manuscript models in which we do find that changes in subjective cognitive complaints predict changes in depression and anxiety severity post-stroke, here we find no evidence that the reverse process is the case (as shown by the lack of interaction effects). * *p* < 0.05, \*\**p* < 0.01, \*\*\**p* <0.001

